# Spatial Transcriptomics Characterisation of Radionecrotic Changes in Glioblastoma Patients

**DOI:** 10.1101/2025.09.25.25336313

**Authors:** Zaira Seferbekova, Michael Ritter, Gleb Ruckhovich, Sophia Schinkewitsch, Nela Köberer, Niklas Grassl, Tobias Kessler, Violaine Goidts, Miriam Ratliff, Christel Herold-Mende, Sandro M. Krieg, Nima Etminan, Michael Platten, Wolfgang Wick, David Reuss, Andreas von Deimling, Felix Sahm, Moritz Gerstung, Abigail K. Suwala

**Author notes:** **Corresponding**: Felix Sahm, Im Neuenheimer Feld 224, 69120 Heidelberg, Moritz Gerstung, Im Neuenheimer Feld 280, 69120 Heidelberg, Abigail Suwala, Im Neuenheimer Feld 224, 69120 Heidelberg.

## Abstract

**Background:** IDH-wildtype Glioblastoma (GB) is the most prevalent primary CNS tumour in adults. The standard treatment involves radiotherapy, which can cause radionecrotic changes. Progressive GB and radionecrotic changes can be challenging to differentiate, as they present with similar symptoms and appear alike on MRI. Histopathological examination remains the gold standard of diagnostics. To this date, little is known about the biological mechanisms underlying radionecrotic changes.

**Methods:** The cohort comprised ten samples from nine patients diagnosed with GB, who underwent first-line standard treatment including surgery, radio- and chemotherapy with temozolomide. Subsequent radiological examination identified tumour progression in all patients, necessitating a second surgery. Based on histopathological examination of the material collected from the second surgery, four patients were diagnosed with tumour recurrence, four manifested with radionecrotic changes, and one patient demonstrated both. The spatial single cell transcriptomics profiling of the samples was conducted using the Xenium platform.

**Results:** We generated a comprehensive spatial single cell transcriptomics atlas of progressive GB and brain tissue with radionecrotic changes. Tumour cells were detected in samples from both groups. Progressive GB samples contained OPC/NPC-like and proliferating tumour cells with high *EGFR* expression. In radionecrotic samples, tumour cells exhibited lower *EGFR* expression even in the presence of gene amplification and did not show proliferation markers. Border-associated macrophages infiltrated the tissue and might have promoted gliosis in radionecrotic samples.

**Conclusions:** This study delineates a complex spatial architecture of brain tissue with post-treatment changes and its discrepancies from progressive GB, facilitating future research into novel treatment strategies.

**Key Points:**

1. A spatial transcriptomics atlas enables comparison of GB and radionecrotic changes.
2. GB contains progenitor and proliferating tumour cells with high *EGFR* expression.
3. Radionecrotic changes are linked to infiltrating BAMs and tumour cells with low *EGFR* levels.

**Importance of the Study:** Radionecrosis is a delayed treatment effect in patients with GB undergoing radiotherapy. Despite its frequent occurrence, very little is known about the underlying biology and cells involved in the process. This study presents the first spatially-resolved single cell atlas of histologically diagnosed radionecrotic changes compared to progressive GB consisting of 1,189,460 cells across ten samples from nine patients. Most importantly, we detect a high number of tumour cells in samples with radionecrotic changes. However, in contrast to progressive GB, these cells have lower *EGFR* expression even in the presence of genetic amplification and do not proliferate. Furthermore, tumour cells colocalise with BAMs that release cytokines, which may promote the formation of gliosis in patients with radionecrotic changes of brain tissue. Our study gives valuable insights into cell-cell interactions in radiation necrosis, which can inform future studies aimed at improving treatment strategies for GB patients.

## Introduction

IDH-wildtype Glioblastoma (GB) represents the most prevalent primary malignant CNS tumour in adults with a high mortality rate^1^. GB is distinguished by three principal histopathological features: a diffuse growth pattern, microvascular proliferation, and necrosis^2^. The necrotic core in GB is surrounded by hypercellular zones with elongated tumour cells, known as pseudopalisades^3^.

The current standard of care therapy for GB involves maximally safe surgical resection, followed by the administration of temozolomide and concomitant radiotherapy^4^. Nevertheless, due to the aggressive infiltrative growth of the tumour and its location, complete surgical removal is not feasible, and GB is prone to rapid recurrence. Furthermore, radiotherapy is commonly associated with complications such as radionecrosis, which occurs in around a fifth of GB patients^5,6^. Radionecrosis manifests from several months to several years following the initial treatment^7^ and is thought to mainly affect non-tumourous brain tissue exposed to radiation. Histological radionecrotic changes encompass coagulative necrosis, hyalination of vessel walls, and severe vasculature damage, which occur in the white matter^8^.

The clinical manifestation of radionecrosis and progressive GB can present similarly, which complicates their radiological differentiation. Although MRI has been utilised for the evaluation of disease status^9^, histopathological examination remains the gold standard for differentiating GB progression from post-treatment complications^10^. However, histologically, progressive GB can also present with reactive changes caused by prior therapy, further complicating the diagnostic process. Nonetheless, the precise clinical diagnosis is imperative, given that progressive GB and radionecrosis necessitate divergent treatment strategies and misdiagnosis may result in worsening of clinical symptoms^4^. Despite considerable efforts, the underlying biology of radionecrosis and how it differs from progressive GB remain poorly understood.

In the present study, we employed spatial single cell transcriptomic profiling of hundreds of genes to generate a comprehensive atlas of progressive GB and radionecrotic changes encompassing several brain-resident cells and tumour cell states. The subcellular spatial resolution enabled the establishment of complex spatial tissue architectures shaped by necrosis in both diagnoses. We found that, in radionecrotic changes, border-associated macrophages infiltrate the necrotic tissue and promote gliosis, and that mutant cells, although prevalent, had low expression of *EGFR*. In contrast, progressive GB was characterised by the presence of OPC/NPC-like and proliferating tumour cells that maintained high *EGFR* expression. We believe that this study offers a deeper understanding of the spatial biology of progressive GB and radionecrosis, paving the way for future research into potential biomarkers and treatment approaches.

## Materials and Methods

### Specimen Collection

We collected formalin-fixed and paraffin-embedded (FFPE) material from nine patients initially diagnosed with GB, IDH-wildtype, WHO grade 4 and undergoing standard of care treatment with temozolomide and radiotherapy (**Supp. Table S1**). At the time of material collection, all patients had MRI findings consistent with tumour progression according to RANO criteria^9^ following initial therapy. Four patients were histologically diagnosed with tumour progression, four demonstrated radionecrotic changes, and one patient presented with both. Necrotic areas were annotated by a neuropathologist on consecutive haematoxylin and eosin (H&E) slides.

### Spatial Single Cell Transcriptomics Experiment

#### Sample preparation

Areas including necrosis and surrounding tissue were selected by a neuropathologist. Respective 5 µm thick tissue sections were floated on a water bath until all wrinkles were eliminated, then carefully mounted onto Xenium slides, ensuring the fiducial frame remained intact. Slides were incubated at 42LJ°C for 10 minutes on a thermocycler using a compatible adapter. After incubation, slides were air-dried and stored overnight in a desiccator.

#### Spatial single cell transcriptomics experiment

The Xenium *In Situ* platform uses a predefined panel for profiling expression of targeted transcripts. We employed the off-the-shelf Xenium Human Brain Panel consisting of 266 genes (PN-1000599). Samples were processed using the Xenium analyzer platform according to manufacturer’s protocols.

### Downstream Spatial Single Cell Transcriptomics Data Analysis

#### Cell segmentation

For whole cell segmentation, Baysor (v0.6.1)^11^ was used. As a prior segmentation we used nuclear segmentation based on DAPI staining. Minimal number of molecules for a cell to be considered as real was set to 5 and confidence of the prior segmentation was set to 0.1. After segmentation, a gene count per cell expression matrix was generated separately for each sample.

#### Data processing and cell annotation

For computational analysis we used Scanpy (v1.10)^12^ and Python (v3.9). First, we filtered out 1,360,821 low quality cells (56%) containing fewer than 30 transcripts in total. The raw counts for the remaining 1,065,178 cells (44%) were normalised, *log1p*-transformed, and scaled to have unit variance and zero mean per gene within each sample. Principal component analysis was performed on the scaled data with default parameters and a neighbourhood graph was computed using 50 principal components and a neighbourhood size of 15. The neighbourhood graph was embedded using uniform manifold approximation and projection (UMAP) with default parameters. The neighbourhood graph was clustered using the Leiden algorithm with resolution 0.5. Each cluster was annotated based on expression of cell type markers (**Fig. 2B**). This initial annotation resulted in identification of six major cell types (Oligodendrocytes, Neurons, Vascular, Myeloid, Lymphoid, and Tumour cells). Counts for Vascular cells were re-scaled and the cluster was re-analysed to annotate Perivascular fibroblasts and Endothelial cells. Similarly, the Neuron cluster was re-analysed to annotate Inhibitory and Excitatory neurons. The procedure was repeated for the Lymphoid cells to annotate T cells. Since 14,185 cells from the Lymphoid cluster did not express any specific cell type markers they were labelled as “Unknown”.

#### Cell annotation refinement

To annotate low-quality filtered out cells and refine produced annotations we used single cell variational inference (scVI) models implemented in scvi-tools (v1.2.0)^13^. To obtain the latent representation, we trained a scVI model^14^ on a random sample from the annotated part of the dataset (115,165 cells stratified by patient and cell type excluding the “Unknown” cluster) with Poisson gene likelihood distribution, 64 nodes per hidden layer, and a 16-dimensional latent space. Cell clustering using the scVI latent space split Endothelial cells into two smaller clusters. One of the two clusters expressed *NR2F2* and *CDH6* and was relabelled as “Pericytes/Vascular smooth muscle cells”. We trained a new scVI model with the same parameters and the new cell type included into the labels. The newly trained scVI model was used to generate decoded transcript fractions in each cell. To transfer annotations to the whole dataset we initialised a scANVI model^15^ with weights from the pretrained scVI model and trained it on the same dataset sample. We used the trained scANVI model to predict cell type probabilities for the whole dataset. Cells with more than 10 total transcripts and cell type probability above 0.8 were assigned the predicted cell type label. Since Perivascular fibroblasts had the lowest model confidence (**Supp. Fig. S1A**), we only assigned this label to cells with more than 30 total transcripts. The remaining cells were labelled “Uninformative cells”.

#### Transcription programs

To identify transcription programs in the Tumour and Myeloid clusters, consensus NMF (v1.6.0)^16^ was used. For the Tumour cluster, ten programs corresponded to the highest stability-to-error ratio (**Supp. Fig. S1B, Supp. Table S2**). Upon closer examination, we discovered that Myeloid and Neuronal programs contributed to less than 20% of total transcriptomic signal in the Tumour cells. Additionally, Invasion and Hypoxic MES2b-like programs were predominantly sample-specific. Consequently, we excluded these four programs from further analysis. For the Myeloid cluster, the optimal stability-to-error ratios were achieved for four and ten programs (**Supp. Fig. S1C**). However, the top contributing genes for most of the ten programs corresponded to markers of previously annotated cell types. Accordingly, the additional programs likely corresponded to noise picked up by factorisation. Therefore, the four-program solution was selected for further analysis of Myeloid cells (**Supp. Table S3**).

#### Spatial community detection

A spatial graph was calculated based on Delaunay triangulation, followed by filtering out neighbours more than 100 µm apart. To identify spatial communities, we next used the graph to compute neighbourhood enrichment by permutation test implemented in Squidpy (v1.5.0)^17^.

#### Overexpressed genes prioritisation

To identify genes that were overexpressed in BAMs in close proximity to Gliosis MES1-like cells, a custom gene prioritisation strategy was implemented. Firstly, the distances from each BAM to the closest Gliosis MES1-like cell were calculated, with the results then being filtered to include only cells closer than 250 µm. Within each sample, a Spearman correlation coefficient was then calculated between the distances and gene transcript fractions in BAMs. Next, to obtain correlation coefficients for each diagnosis, the median was calculated for samples from either group. Finally, genes exhibiting a correlation coefficient smaller than the 0.05 quantile of the diagnosis-level distribution were considered overexpressed in BAMs in closer proximity to Gliosis MES1-like cells. Similarly, genes with a correlation coefficient larger than the 0.95 quantile were considered overexpressed in BAMs further away from Gliosis MES1-like cells. The same procedure was repeated for Gliosis MES1-like cells in relation to BAMs and for all Tumour cells in relation to necrotic areas, with 100 µm used as a maximum distance threshold for the former and no threshold used for the latter.

### Immunohistochemistry

Immunohistochemistry (IHC) staining was performed on a Ventana BenchMark ULTRA Immunostainer (Ventana Medical Systems, Tucson, USA) on FFPE sections. Stainings for CD163 and Ki67 were performed on 0.5 µm thick sections (CD163, clone MRQ-26, Roche; Ki67, clone MIB-1, dilution 1:100, Dako Agilent, Santa Clara, CA, USA). Stained slides were scanned on the Aperio AT2 Scanner (Aperio Technologies, Vista, USA) and digitalised using Aperio ImageScope software (v12.3.2.8013).

### Copy Number Variation Detection

For DNA methylation, the Infinium MethylationEPIC (v2.0, Illumina, San Diego, USA) was used according to manufacturer’s instructions. The data were processed as previously described^18^. Copy Number Variation (CNV) profiles were derived from IDAT files using the R/Bioconductor package conumee^19^, following additional baseline correction based on the B allele frequency of single nucleotide polymorphisms included on the BeadChip for DNA fingerprinting^20^.

### Variant Allele Frequency Measurement

DNA was extracted and enriched using Agilent SureSelect technology with a targeted panel of 201 tumour-relevant genes, as previously described^21^. Sequencing was conducted on the Illumina NovaSeq X platform following the manufacturer’s protocol.

## Results

### Mutant Cells Are Prevalent in Samples with Radionecrotic Changes

In our diagnostic routine, we received tissue from two patients with clinical history of GB. Both patients underwent first-line treatment including surgical resection, radio-and chemotherapy with temozolomide. After initial treatment, both patients presented with lesions suspicious for tumour progression in MRI scans. Second surgery was performed, and the tissue was sent for neuropathological examination.

The sample obtained from the first patient exhibited histological criteria for GB, including high cellularity of pleomorphic cells, mitotic activity, microvascular proliferation and pseudopalisading necrosis (**Fig. 1A**, *top*). The results of IHC staining revealed the presence of densely packed populations of Ki67-positive cells (**Fig. 1B**, *top*). Therefore, the patient was diagnosed with progressive GB. In the sample from the second patient, a low number of Ki67-positive cells were dispersed across the sample (**Fig. 1B**, *bottom*), and the characteristic pseudopalisades, unequivocal tumour cells, and mitotic figures were absent (**Fig. 1A**, *bottom*). Instead, radionecrotic changes, namely, coagulative necrosis, reactive astrocytes, and thickened blood vessels were present.

**Figure 1.**
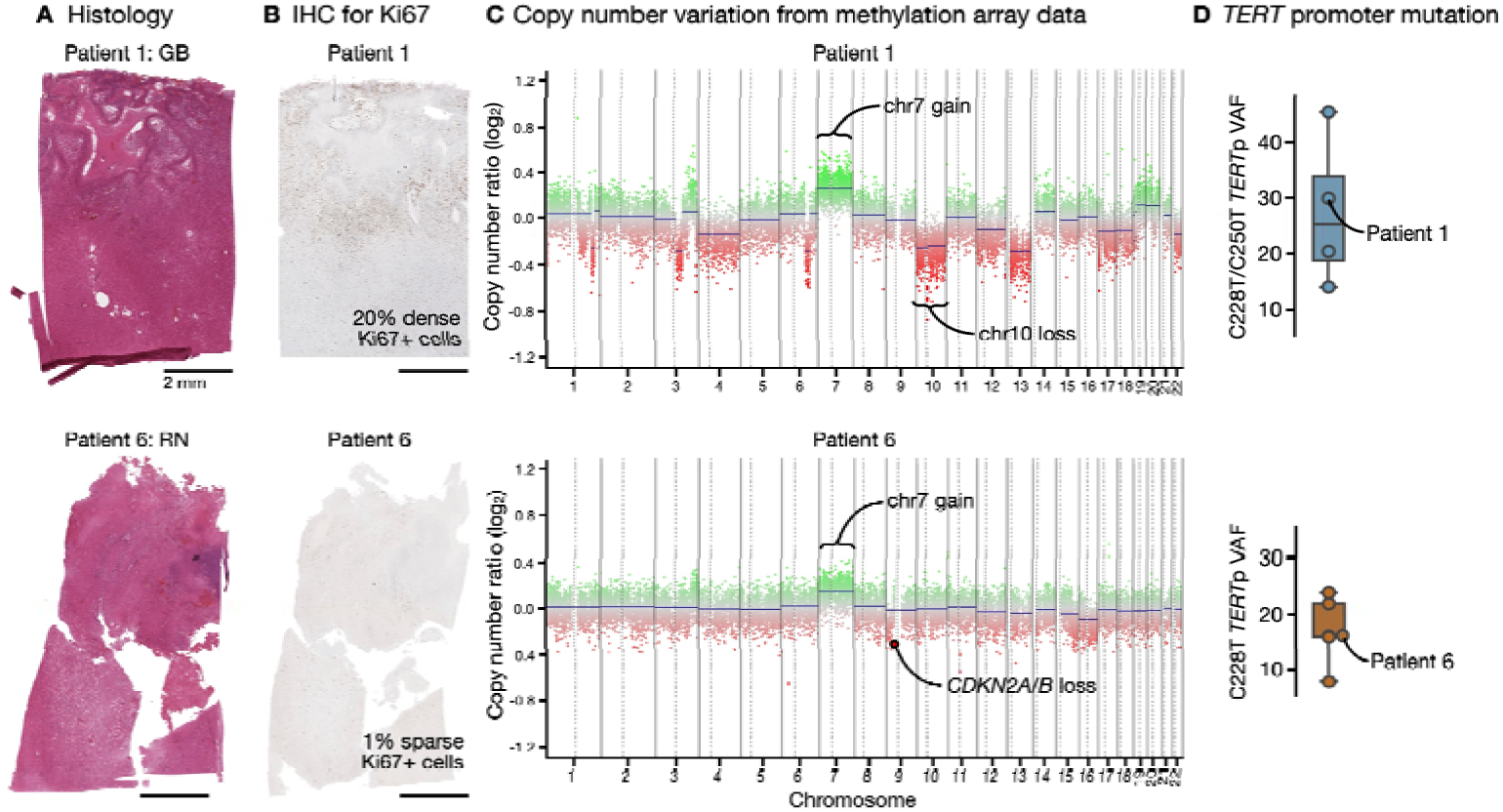
Tumour cells are found in patients with progressive GB and radionecrotic changes. *(A)* H&E whole slide images of samples from two patients with progressive GB (*top*) and radionecrotic changes (*bottom*). *(B)* Staining for a proliferation marker Ki67 is positive in progressive GB (*top*) and radionecrotic changes (*bottom*). *(C)* CNV analysis identifies hallmark copy number events and mutations in both patients. *(D)* Cells with hallmark mutations found in patients with progressive GB (*top*) and radionecrotic changes (*bottom*). GB, glioblastoma; IHC, immunohistochemistry; RN, radionecrotic; *TERT*p, *TERT* promoter; VAF, variant allele frequency.

Subsequent analysis of DNA methylation array data revealed GB hallmark events in bulk CNV profiles of both patients (**Fig. 1C**), a finding that was unexpected. To assess whether this was a recurring phenomenon, a cohort of 10 samples was assembled (**Table 1, Supp. Table S1**). The cohort comprised nine patients treated in accordance with the standard of care protocol. After initial treatment, four patients were histologically diagnosed with progressive GB, four exhibited histological radionecrotic changes of brain tissue, and one patient presented with both. We evaluated C228T and C250T variant allele frequencies in the *TERT* promoter region, as *TERT* promoter mutations are a hallmark of GB^22^. The analysis demonstrated that mutant cells were present in samples from both groups (**Fig 1D**).

**Table 1.** Molecular data for cohort patients.

| Patient | Histology | *MGMT*p status | *TERT*p status | *TERT*p mutation VAF (%) | Ki67 (%) | *EGFR* status | *CDKN2A/B* status |
| --- | --- | --- | --- | --- | --- | --- | --- |
| Patient 1 | GB | Unmethylated | C228T | 29.95 | 20 | Balanced | Balanced |
| Patient 2 | GB | Unmethylated | C228T | 20.39 | 20 | Amplified | Homozygous |
| Patient 3 | GB | Methylated | C250T | 13.91 | 15 | Balanced | Homozygous |
| Patient 4 | GB | Methylated | Wild type | Wild type | 6 | Balanced | Homozygous |
| Patient 5 | RN | Unmethylated | C228T | 21.96 | 3 | Amplified | Hemizygous |
| Patient 6 | RN | Methylated | C228T | 16.03 | 1 | Balanced | Hemizygous |
| Patient 7 | RN | Methylated | C228T | 15.88 | 3 | Balanced | Hemizygous |
| Patient 8 | RN | Methylated | C228T | 23.87 | 1 | Balanced | Homozygous |
| Patient 9 | GB | Unmethylated | C228T | 45.43 | 50 | Amplified | Homozygous |
|  | RN |  | C228T | 7.69 | 5 |  |  |

As all samples exhibited molecular signals indicative of GB, we deemed it prudent to refer to the samples according to their histological characteristics in order to avoid ambiguity. Consequently, we employed “GB histology” to denote samples with histological evidence of GB. The remaining samples, which exhibited radionecrotic changes histologically, were grouped under the designation “Radionecrotic histology”.

In summary, the collective evidence, derived from molecular data, demonstrated that samples with radionecrotic histology contain numbers of tumour cells comparable to those in samples with GB histology. This finding prompted a thorough comparison of the two histologies, with the aim of characterising the tumour cells and spatial communities they may form with brain-resident cells.

### A Spatially-Resolved Transcriptomics Atlas of Radionecrotic and GB histology

To investigate the spatial cellular landscape across the two histologies, samples from the assembled cohort were processed using the Xenium *In Situ* platform and the off-the-shelf Xenium Human Brain Panel (**Fig. 2A**). Following segmentation and cell annotation (see **Materials & Methods**), the final spatial single cell transcriptomics dataset comprised 1,189,460 cells with 112,423,484 transcripts. A total of eight tumour microenvironment (TME) cell types in addition to Tumour cells were identified employing a set of previously established literature markers (**Fig. 2B-C**).

**Figure 2.**
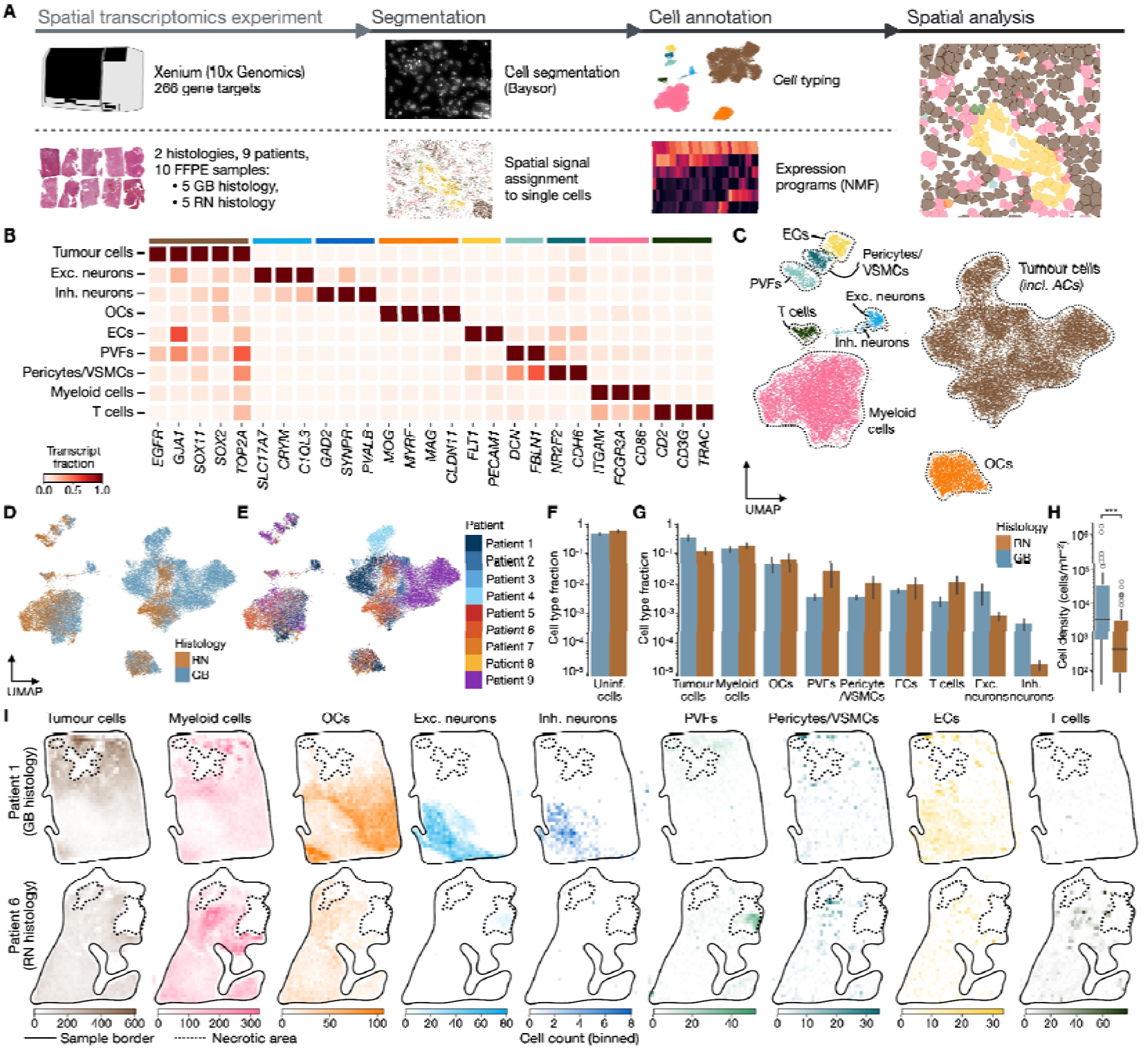
Cellular composition of GB and radionecrotic samples. *(A)* Experimental and analytical pipeline of spatial characterisation of the two sample groups. *(B)* Marker genes used for cell annotation. UMAP coloured by nine major cell types *(C)*, histology *(D)*, and patient ID *(E)*. Normalised cell proportions for filtered out *(F)* and kept *(G)* cells coloured by histology. *(H)* Comparison of cell densities between the groups. ***, *P* < 0.001, two-sided Mann-Whitney *U* test. *(I)* Binned spatial distribution of cell types for two selected GB *(top)* and radionecrotic *(bottom)* samples. AC, astrocyte; EC, endothelial cell; exc., excitatory; FFPE, formalin-fixed paraffin-embedded; inh., inhibitory; NMF, non-negative matrix factorisation; OC, oligodendrocyte; PVF, perivascular fibroblast; VSMC, vascular smooth muscle cell.

It is important to note that one large cluster was characterised by expression of tumour and astrocytic markers (**Supp. Fig. S2**). Furthermore, no separate cluster corresponding to brain-resident astrocytes was observed. Given that GB contains tumour cell populations that are transcriptionally similar to astrocytes^2^, and due to the number of genes being too limited to perform CNV analysis, it was hypothesised that the cluster contained a mixture of brain-resident and malignant cells. Subsequently, the cluster was annotated as “Tumour cells (including astrocytes)”. Interestingly, a comparison of Tumour cell fractions from the spatial single cell transcriptomics data and the expected numbers based on molecular signal suggested that the current annotation may underestimate the amount of GB cells (**Supp. Fig. S2-3**). Similarly, we refrained from establishing a rigid distinction between macrophages and brain-resident microglia within the Myeloid cluster, as myeloid populations in GB constitute a continuum rather than falling into distinct, transcriptionally-defined cell groups^23^. Both the Tumour and Myeloid clusters were later reviewed to identify transcription programs that are characteristic of various cell states (see below).

The eight annotated TME cell types and Tumour cells were shared among patients with both GB and radionecrotic histologies (**Fig. 2D-E**). The two groups did not differ significantly in the fraction of cells containing less than 30 transcripts in total (**Fig. 2F**, *P* > 0.05).

Moreover, we did not observe significant differences in any of the annotated cell type fractions between GB and radionecrotic histologies (**Fig. 2G**, *P* > 0.05, two-sided Mann-Whitney *U* test with Benjamini-Hochberg correction). Among the nine cell types, the most abundant one was Tumour cells, followed by Myeloid cells and Oligodendrocytes, regardless of the group. Neurons were the rarest cell type. Finally, samples with GB histology exhibited higher cell densities compared with radionecrotic histology (**Fig. 2H**, *P* = 1.29×10^-5^, two-sided Mann-Whitney *U* test).

We next explored the spatial distribution of the annotated cell types (**Fig 2I**, **Supp. Fig. S4**). All samples exhibited a dispersed distribution of Oligodendrocytes, Tumour cells, and Myeloid cells, without any discernable spatial patterns. Endothelial cells, Pericytes, and T cells were found to colocalise with each other and to be scattered across the entire sample area. Neurons were predominantly present in samples with GB histology and localised in a confined spatial region. This region represented grey matter, wherein neuronal somas with the majority of transcript molecules are located and may be captured during the experiment. By contrast, radionecrotic changes are primarily observed in the white matter, which is largely composed of axons^24^. It is conceivable that the number of transcript molecules present within axons is insufficient for effective capture.

In summary, a comprehensive spatial single cell transcriptomic atlas encompassing ten samples from nine patients presenting with GB and radionecrotic histologies was constructed. The atlas facilitated a detailed comparative analysis of the two groups.

### Tumour Cell *EGFR* Expression Is Lower in Radionecrotic Samples

To investigate the expression diversity present within the Tumour cluster, ten transcription programs were identified for Tumour cells employing consensus NMF^16^ (**Fig. 3**, **Supp. Table S2**, see **Materials and Methods**). The annotation of these programs was informed by the findings of earlier studies that examined transcription cell states in GB^2,25,26^. A total of six programs were selected for further analysis, given that two programs were sample-specific and two others contributed to less than 20% of the total transcriptomic signal in Tumour cells (**Fig. 3A**, **Supp. Fig. S5**, see **Materials and Methods**).

**Figure 3.**
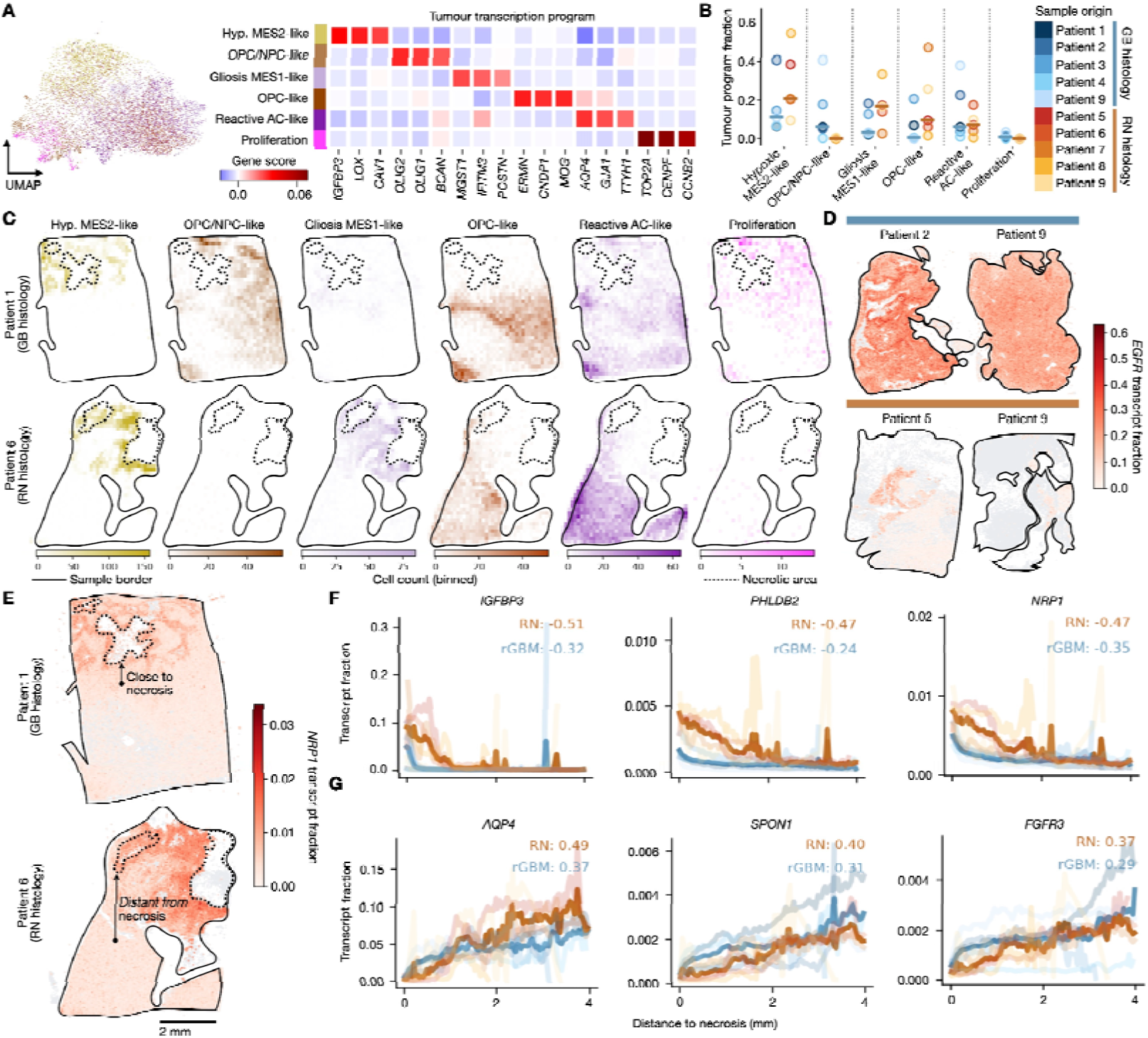
Tumour cells are characterised by ten distinct expression programs. *(A)* Annotation of the tumour transcription programs. *(B)* Tumour program fractions coloured by sample. *(C)* Binned spatial distribution of tumour transcription programs for an exemplary GB *(top)* and radionecrotic *(bottom)* samples. *(D) EGFR* expression in three patients with *EGFR* amplification. *(E)* Cell distance to necrosis can correlate with gene expression. *(F)* Genes with higher expression in tumour cells close to necrosis and *(G)* distant from necrosis. Numbers correspond to Spearman correlation coefficients between tumour cell distance to necrosis and tumour gene expression. Hyp., hypoxia; prolif., proliferation.

Four tumour transcription programs corresponded to established transcription GB states, namely mesenchymal (MES), oligodendrocyte progenitor (OPC), and neural progenitor (NPC)-like states (**Fig. 3A**). The Hypoxic MES2-like cells constituted the predominant population within the Tumour cluster (**Fig. 3B**). Spatially, Hypoxic MES2-like tumour cells were discovered perinecrotically (**Fig. 3C**, **Supp. Fig. S6A**), suggesting that the program may primarily represent a response to hypoxic conditions. The OPC/NPC-like program exhibited both progenitor (*OLIG2, OLIG1, SOX4*) and stem (*BCAN, PTPRZ1, NOTCH1*) cell markers. Notably, the OPC/NPC-like cells were found to be less prevalent in radionecrotic compared to GB samples, although the difference did not reach statistical significance (*P* > 0.05, two-sided Mann-Whitney *U* test with Benjamini-Hochberg correction). When present, OPC/NPC-like cells populated spatial areas away from the necrotic core. Gliosis MES1-like program was characterised by both mesenchymal and astrocytic markers (*IFITM3, POSTN, MGST1, THBS1*). Spatially, Gliosis MES1-like cells were observed in regions adjacent to Hypoxic MES2-like cells but at a greater distance from necrosis. The OPC-like program was enriched in oligodendrocytic (*CNDP1*, *ERMN*) and OPC (*MAG*) markers. The Reactive astrocyte-like (AC-like) program was characterised by *AQP4*, *FGFR3*, *SOX9*, and *SPON1*, markers specific to brain-resident astrocytes.

It is interesting to note that none of the transcription programs was uniquely associated with *EGFR*, a gene that is thought to be a key driver in GB^27^. The present study comprised three patients with *EGFR* amplification: one patient with GB histology, one with radionecrotic histology, and one with both observed. Evidently, although transcription programs were mostly shared between the two sample groups, the expression levels of *EGFR* in radionecrotic samples were lower in comparison to GB samples (**Fig. 3D**), a finding which was particularly striking for two samples from the same patient. The expression difference could not be attributed to genetic loss of the *EGFR* amplification (**Supp. Table S4**) and therefore was more likely a result of *EGFR* downregulation in samples with radionecrotic histology.This finding is of interest given that downregulation of a key driver gene in tumour cells following treatment exposure may have clinical significance.

Finally, we assessed how distance to necrosis may influence tumour cell expression. Interestingly, Tumour cells in closer proximity to necrotic regions exhibited elevated expression of genes associated with cell locomotion (*NRP1*, *IGFBP3*, *PHLDB2*), irrespective of their transcription program (**Fig. 3E-F**, see **Materials and Methods**). It has been previously demonstrated that pseudopalisading GB cells constitute an actively migrating population^28^. Our present finding suggests that this conclusion could be extended to perinecrotic cells in radionecrotic samples. Finally, Tumour cells located farther away from necrotic regions expressed high levels of astrocytic markers (**Fig. 3G**).

Collectively, we found OPC/NPC-like and Proliferating tumour cells only in GB samples. Additionally, tumour cells in samples with radionecrotic histology expressed lower levels of *EGFR* when compared to GB samples, and that difference could not be explained by the loss of the gene amplification.

### Border-Associated Macrophages Infiltrate Brain Parenchyma in Radionecrotic Samples

In order to characterise myeloid populations with greater detail, consensus NMF was utilised to identify four expression programs within the Myeloid cluster (**Fig. 4**, **Supp. Table S3**, see **Materials and Methods**). This approach enabled the characterisation of brain-resident Microglia that expressed *P2RY12*, *CX3CR1*, and *GPR34* and Border-associated macrophages (BAMs) expressing *LYVE1*, *THBS1*, and *CD163* (**Fig. 4A**). Furthermore, the analysis identified Proliferation and Hypoxia programs, analogous to those observed for Tumour cells.

**Figure 4.**
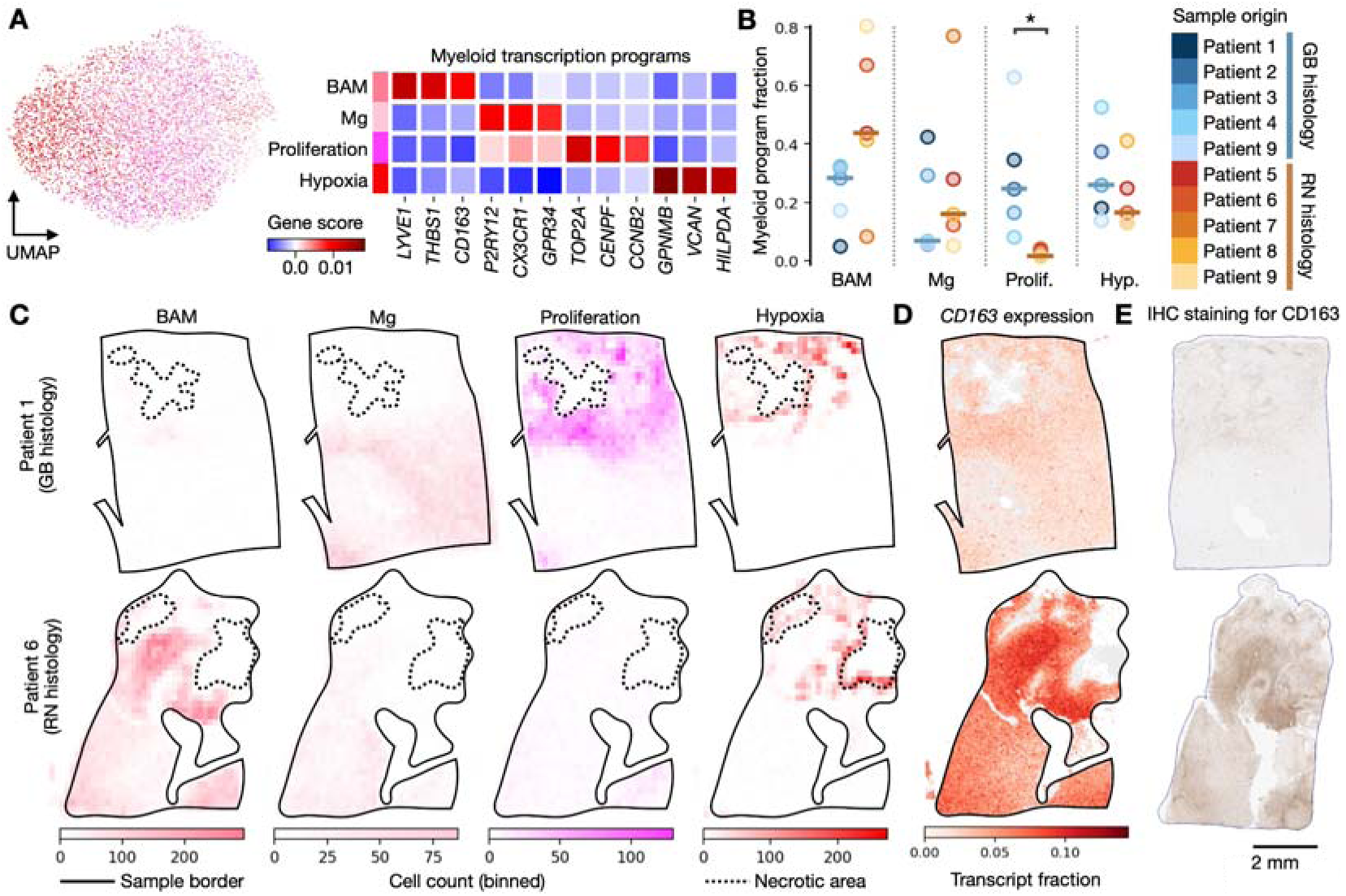
Myeloid cluster consists of four myeloid cell states. *(A)* Annotation of the myeloid transcription programs. *(B)* Myeloid program fractions coloured by sample. *, *P* < 0.05, two-sided Mann-Whitney *U* test with Benjamini-Hochberg correction. *(C)* Binned spatial distribution of myeloid transcription programs for two selected GB *(top)* and radionecrotic *(bottom)* samples. *(D)* BAM marker *CD163* expression in tumour cells in the GB *(top)* and radionecrotic *(bottom)* samples. *(E)* IHC staining for CD163 in the GB *(top)* and radionecrotic *(bottom)* samples. BAM, border-associated macrophage; Mg, microglia.

Samples with GB histology contained significantly greater fractions of Proliferating myeloid cells when compared with radionecrotic samples (**Fig. 4B**, *P* value = 0.0317, two-sided Mann-Whitney *U* test with Benjamini-Hochberg correction). While the differences observed for the three other programs did not reach statistical significance, BAM fractions in four out of five samples with radionecrotic histology exceeded those of all GB samples (**Fig. 4B**). In general, in radionecrotic samples BAMs constituted the most prevalent myeloid population.

We next examined the spatial distributions of the myeloid programs (**Fig. 4C**, **Supp. Fig. S6B**). Similarly to the Hypoxic MES2-like tumour cells, the Hypoxic myeloid cells populated perinecrotic regions in samples with both histologies. In contrast, Proliferating myeloid cells populated areas away from necrotic regions. In the case of BAMs, the location of the cells differed between radionecrotic and GB samples. In GB, BAMs were observed in close proximity to vessels. Conversely, in samples with radionecrotic histology, BAMs were found to diffusely infiltrate the entirety of tissue. To corroborate the finding that BAM signature is expressed outside of the perivascular niche in radionecrotic samples, we compared myeloid single cell *CD163* signal (**Fig. 4D**) and IHC staining for CD163 (**Fig. 4E**). Evidently, the radionecrotic sample demonstrated pronounced positive staining encompassing considerable areas, while the GB sample exhibited staining that was confined to smaller perivascular regions.

The present findings demonstrate that the Proliferation myeloid program is predominantly present in samples with GB histology. Furthermore, in cases of radionecrotic samples, Myeloid cells in the brain parenchyma express the BAM program. This is in contrast to GB histology, wherein BAMs primarily populate perivascular areas.

### Stromal Border-Associated Macrophages Promote Gliosis in Radionecrotic Samples

The final objective of this study was to explore the presence of spatial patterns in samples with GB and radionecrotic histologies (**Fig. 5**). Using permutation, we proceeded to assess whether specific cell groups exhibited a tendency to colocalise with one another, thereby forming recurrent spatial communities (**Fig. 5A-B**, **Supp. Fig. S7**, see **Materials and Methods**).

**Figure 5.**
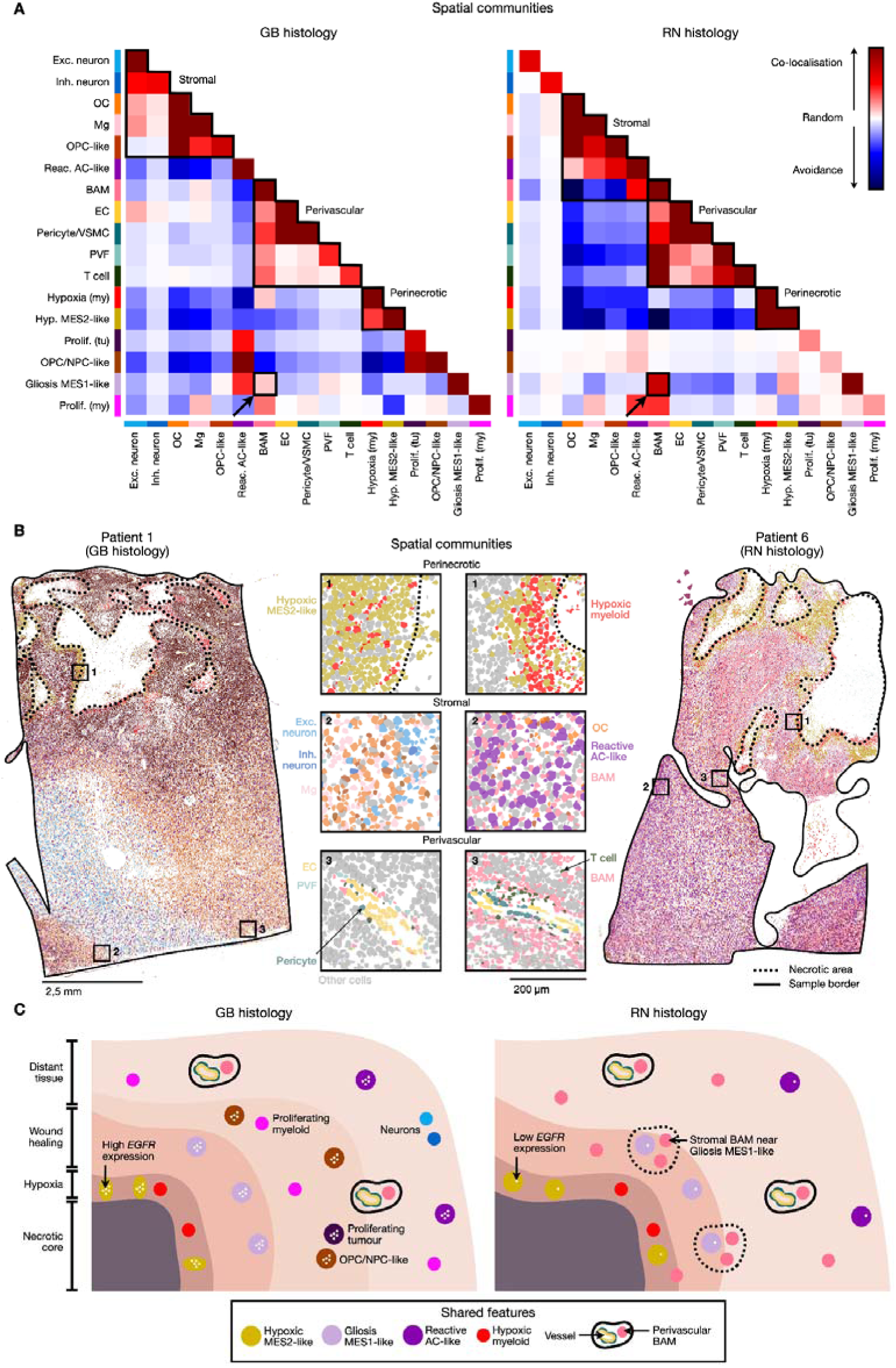
Complex spatial architecture of GB and radionecrotic samples. *(A)* Cooccurring TME cell types and tumour programs form spatial communities in GB *(left)* and radionecrotic *(right)* samples. My, myeloid transcription program; tu, tumour transcription program. *(B)* Spatial distribution of cells within the three communities from *(A)* in two GB *(left)* and radionecrotic *(right)* samples. *(C)* Schematic representation of the reconstructed spatial ecosystems.

As anticipated, Hypoxic myeloid and MES2-like tumour cells colocalised next to necrotic regions, forming a demarcated perinecrotic spatial community reacting to hypoxic stress that was shared between both groups. Furthermore, in GB samples, Proliferating tumour cells colocalised with OPC/NPC-like cells in regions distant to necrosis.

Brain-resident cell types such as Neurons, Oligodendrocytes, and Microglia colocalised in a community we have defined as “Stromal”. The presence of Neurons within the community was only observed in samples with GB histology, which could be attributed to generally lower prevalence of Neurons in radionecrotic samples. Conversely, only in samples with radionecrotic histology Reactive AC-like cells and BAMs were found to colocalise together within the Stromal community.

BAMs, Pericytes, Endothelial, and T cells formed a third spatial community that exhibited a uniform composition in both GB and radionecrotic samples and was annotated as “Perivascular”. This observation suggests that the perivascular niche persists within radionecrotic histology, despite extensive vessel damage.

In accordance with the observations discussed previously, BAMs were present within both the Stromal and Perivascular communities exclusively in samples with radionecrotic histology. Moreover, BAMs were observed to recurrently colocalise with Gliosis MES1-like tumour cells, and this pattern was more prominent in radionecrotic samples (**Fig. 5A**, *see arrow*). The proximity of BAMs to Gliosis MES1-like cells was associated with increased expression of genes that upregulate cytokine production, TNF in particular (**Supp. Fig. S8**), which is known to contribute to gliosis^29^. With regard to BAMs that colocalised with Reactive AC-like cells, such enrichment in genes associated with cytokine production was not observed.

The present study suggests that necrotic tissue associated with GB exhibits a complex spatial organisation, with variations in its characteristics depending on the exact histological diagnosis (**Fig. 5C**). In samples with radionecrotic histology, we observed the colocalisation of BAMs with both Reactive AC-like cells and Gliosis MES1-like tumour cells, accompanied by the upregulation of cytokine production in the vicinity of the latter.

## Discussion

Here, we present the first study giving comprehensive insights into radionecrotic histology compared to GB histology using spatial single cell transcriptomics. We revealed *TERT* promoter mutant tumour cells and CNVs in tissue histologically diagnosed as radionecrosis. Furthermore, samples with radionecrotic histology exhibited high numbers of BAMs infiltrating the entire necrotic tissue and colocalising with Reactive AC-like and Gliosis MES1-like cells. In contrast, GB histology was characterised by the presence and colocalisation of Proliferating and OPC/NPC-like tumour cells. Finally, Tumour cells in GB samples from patients with *EGFR* amplification expressed high levels of *EGFR*, whereas in radionecrotic samples *EGFR* expression was markedly low.

As demonstrated in previous studies, radiation injury is associated with irreversible damage to the NPC and OPC compartments^24^. However, all patients were administered radiotherapy, and radionecrosis typically presents months or years subsequent to that treatment. Consequently, the observed differences in the amounts of OPC/NPC-like cells and *EGFR* expression cannot be solely attributed to exposure to radiation. Instead, we speculate that the discrepancies found in the Tumour cell populations are the result of different environmental pressures acting in the necrotic environment of samples with GB and radionecrotic histologies. It is of interest as such plasticity may potentially allow all tumour cells, rather than merely a select few that survived a selective bottleneck, to enter a state of quiescence and give rise to a new tumour cell population over time.

Similar to a recent study^30^, we did not observe a clear association between *EGFR* amplification and one specific transcription program. Nevertheless, *EGFR* was among the genes that contributed to the OPC/NPC-like program, a program that was predominantly identified in GB histology cases. Moreover, Proliferating tumour cells were found exclusively in GB histology and in close proximity to OPC/NPC-like tumour cells. In postnatal rats, *EGFR* expression has been demonstrated to maintain progenitor cells in a proliferative state^31^. Consequently, it can be speculated that, in GB, the OPC/NPC-like cells that express high levels of *EGFR* may give rise to Proliferating tumour cells.

Finally, in samples with GB histology, BAMs were found to be restricted to perivascular spaces. By contrast, in radionecrotic samples, BAMs were observed to infiltrate the entire brain parenchyma in greater numbers. Furthermore, in parenchyma, these BAMs colocalised with Reactive AC-like and Gliosis MES1-like tumour cells and upregulated the release of cytokines, including TNF, in close proximity to the latter. TNF is associated with therapy resistance in GB^32^ and recognised as a primary inducer of gliosis, a process implicated in neuroinflammation, oedema and glial scar formation^29^. It has been previously established that, in transgenic rodent models of GB, glial scar areas can act as a protective barrier for glioma cells, thereby promoting their survival after therapeutic intervention^33^.

Furthermore, dysregulated cytokines have been linked to neuroinflammation and multiple neurological disorders^34^. The release of cytokines may therefore provide a plausible explanation of symptoms such as cognitive impairment and epileptic seizures that frequently occur in patients with radionecrosis^5^.

A potential limitation of the study is that the second surgery, which enabled material collection, was performed solely on the basis of suspected tumour progression indicated by MRI scans. Consequently, according to imaging, all samples in this cohort satisfied the RANO criteria for tumour progression^9^, with no cases diagnosed with radionecrosis. This may represent an inherent limitation of this study, since it is not possible to ascertain whether the findings can be directly extrapolated on cases of radionecrosis diagnosed solely on the basis of MRI scans. Nonetheless, while the term ‘radionecrosis’ is predominantly employed within the domain of imaging, histology remains the gold standard for confirming the initial diagnosis^10^.

In conclusion, the present findings indicate that the intrinsic spatial properties of necrotic tissue differ between samples with GB and radionecrotic histologies. Specifically, the characteristics of samples with radionecrotic histology include the presence of tumour cells with low *EGFR*, as well as the infiltration of great numbers of BAMs into necrotic brain tissue, the promotion of gliosis, and the upregulation of cytokine production. Conversely, cases with GB histology are distinguished by the presence of progenitor-like and proliferating tumour cells maintaining high *EGFR* expression. Our study gives valuable insights into the spatial biology of tissue with radionecrotic changes and provides a foundation for further research improving the clinical management of patients with GB.

## Required Statements

### Authorship

A.K.S., M.G., and F.S. conceptualised and supervised the study. Z.S. analysed Xenium data, interpreted the results, and prepared figures. S.S., N.K., N.G., T.K., M.Ra., S.K., N.E., M.P., W.W., and F.S. provided patient samples and clinical information. A.K.S. coordinated tissue sample collection, prepared the cohort, and led histopathological annotation of the collected samples. M.Ri. conducted sample preparation for the Xenium experiment. G.R. performed whole cell segmentation. All authors contributed to writing the manuscript.

### Data Availability

Data and code used in this study are available upon reasonable request.

## Acknowledgements

We would like to express our gratitude to the technical staff at the Neuropathology department for their invaluable aid and support. In addition, we would like to acknowledge the excellent assistance provided by the Single-cell Open Lab (DKFZ) in the execution of the single cell spatial transcriptomics experiment.

