## Supplementary Figures for "Spatial Transcriptomics Characterisation of Radionecrotic Changes in Glioblastoma Patients"

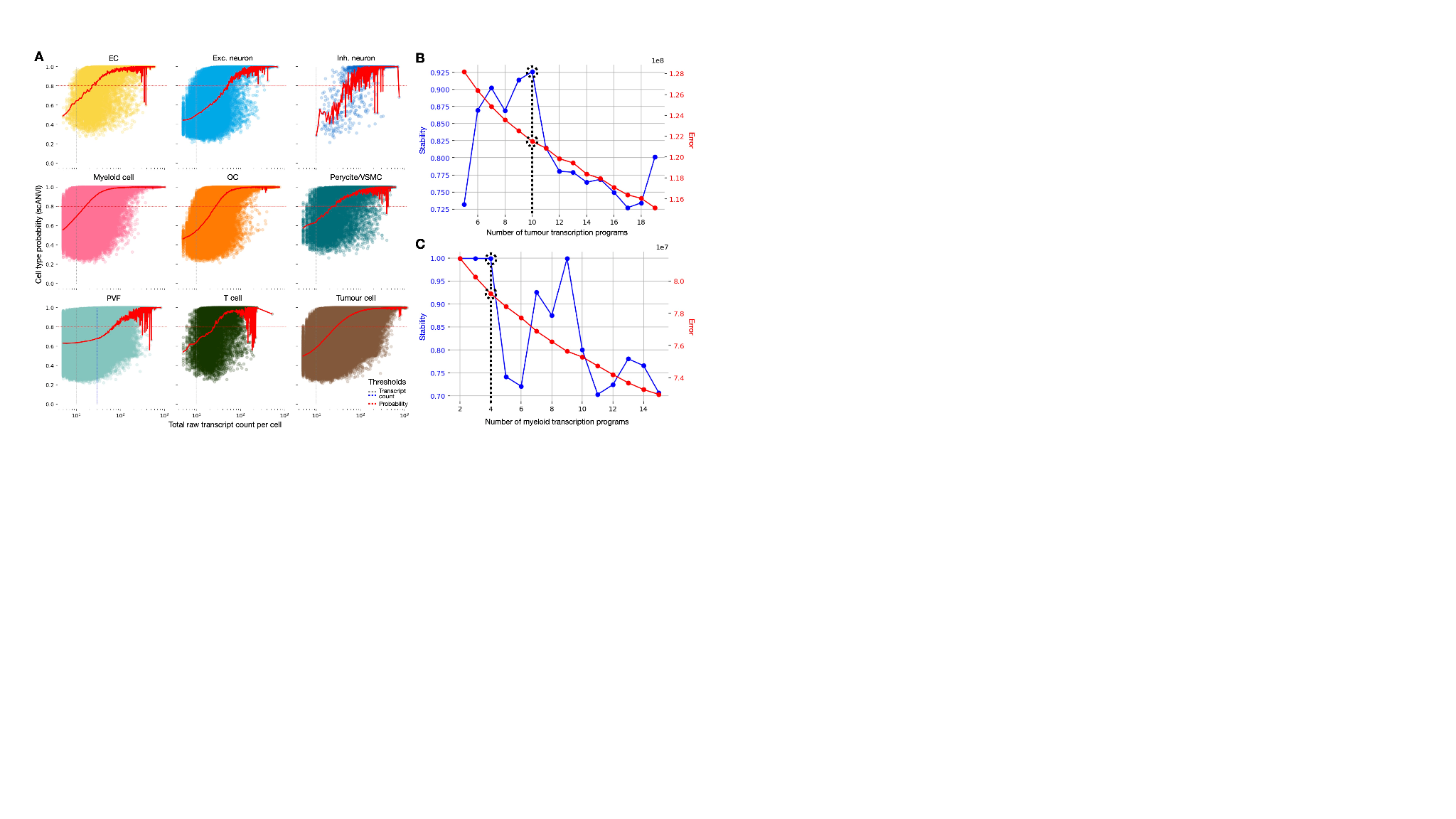

**Supplementary figure S1**. *(A)* Cell type probability for scANVI was generally lower for perivascular fibroblasts. Ten tumour *(B)* and four myeloid *(C)* NMF components corresponded to the optimal solution for consensus NMF.

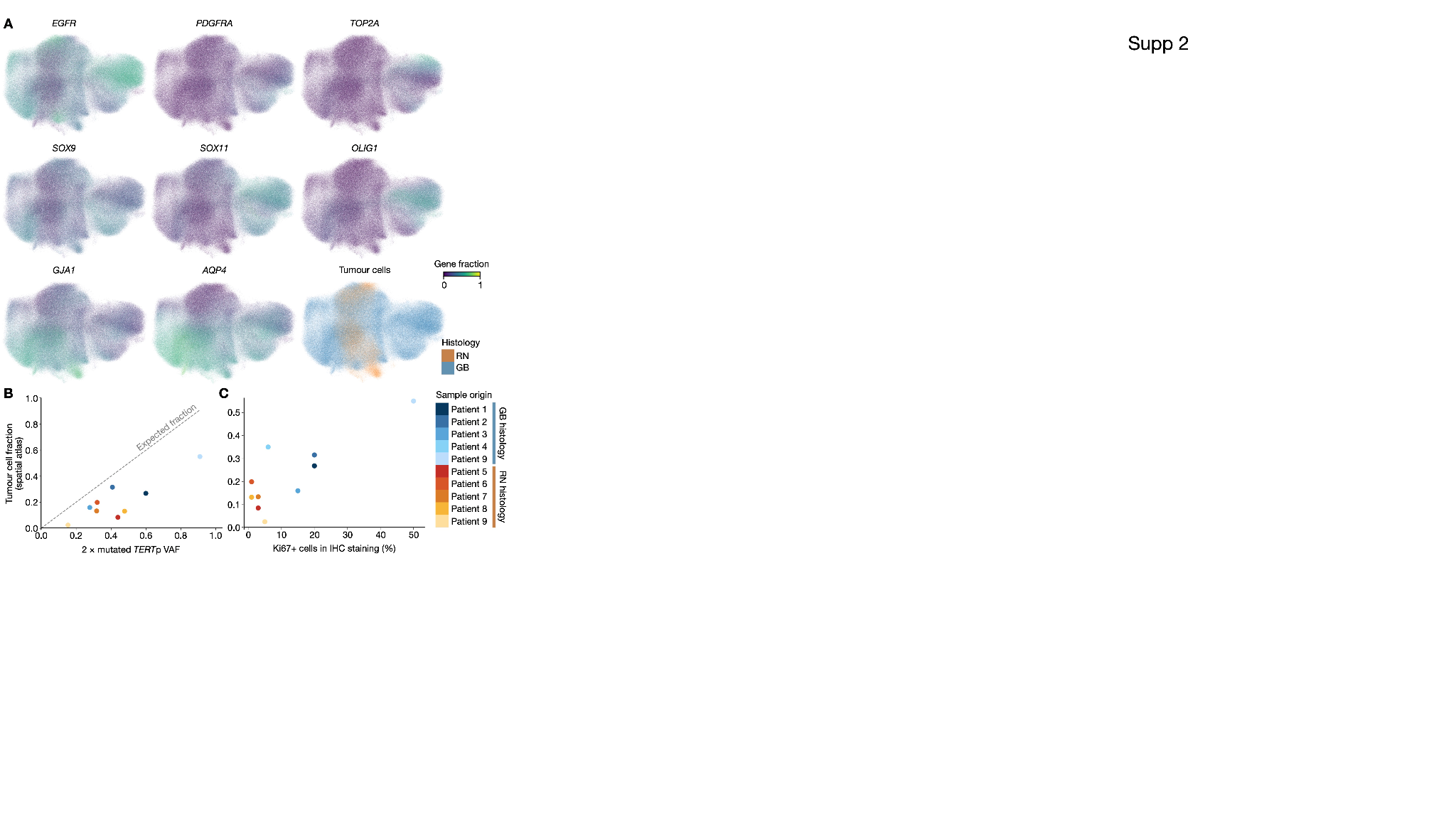

**Supplementary figure S2**. *(A)* Cells in the tumour cluster expressed tumour and astrocyte markers regardless of the patient diagnosis. *(B)* Tumour cell fraction in the spatial single-cell transcriptomics atlas was lower than expected according to mutated *TERT*p VAFs. *(C)* Samples with radionecrotic histology exhibited weak IHC staining for Ki67, whereas GB samples had strong positive staining.

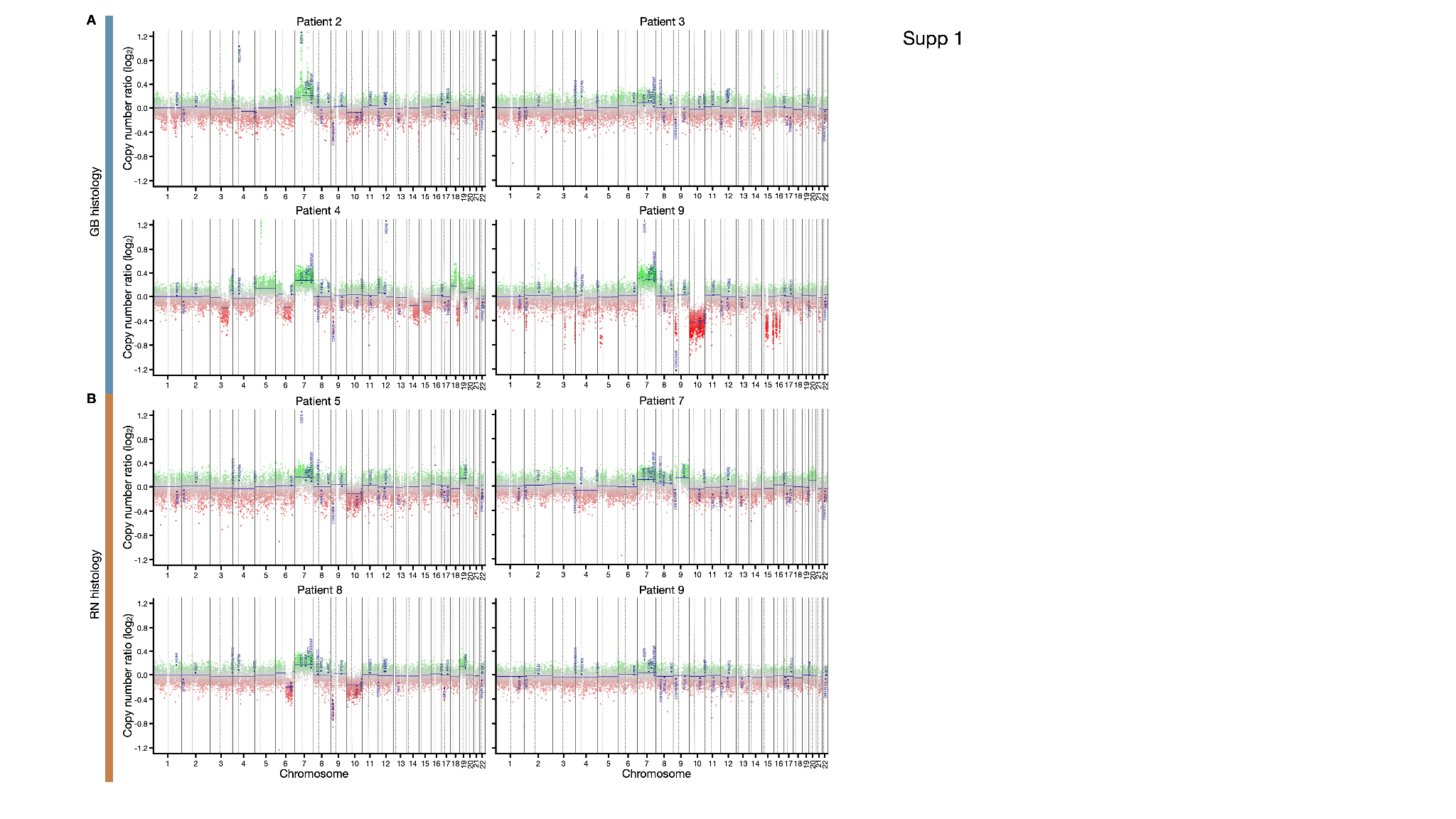

**Supplementary figure S3**. Hallmark CNV events in samples with GB *(A)* and radionecrotic *(B)* histology.

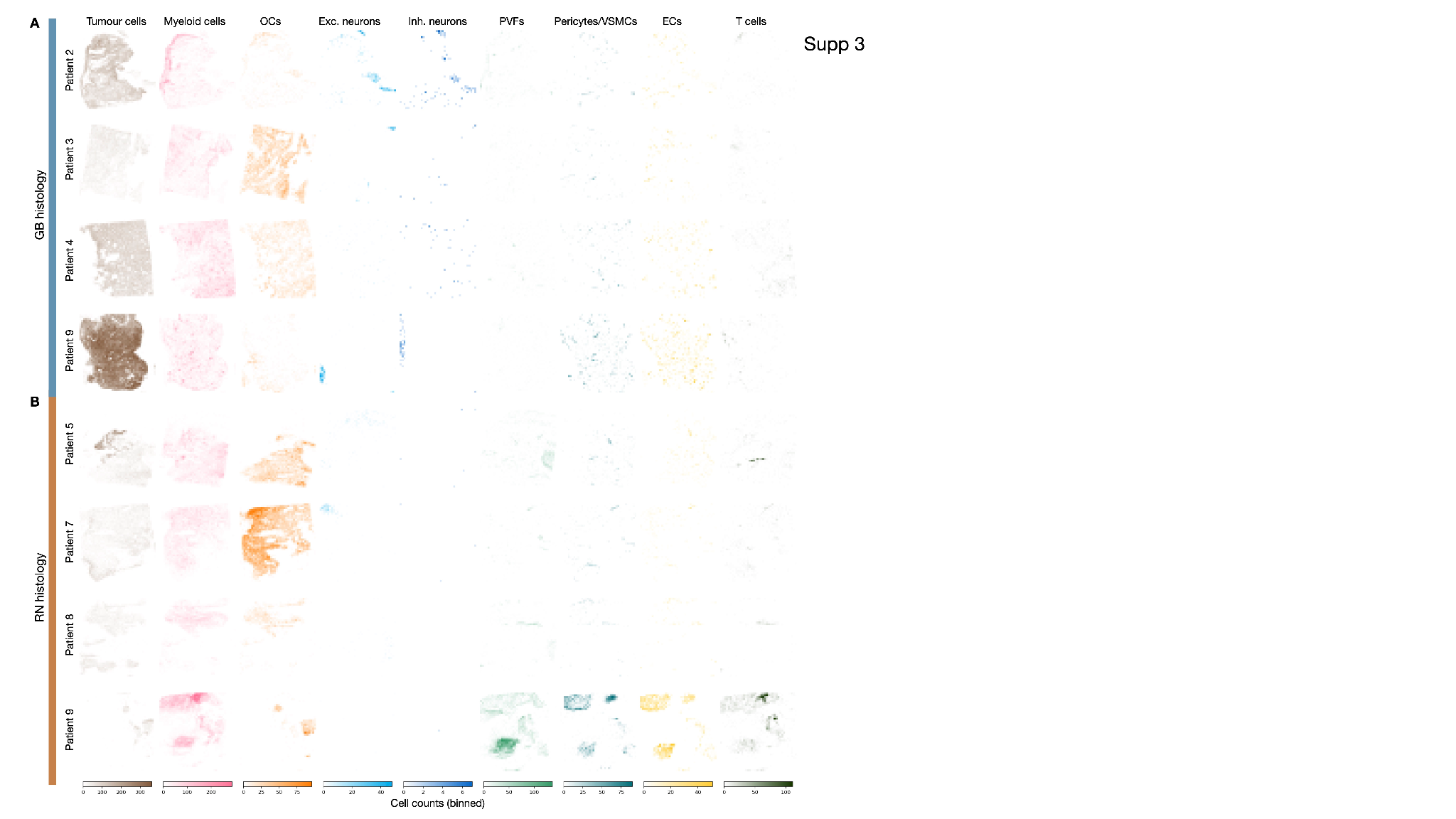

**Supplementary figure S4**. Spatial distribution of cell types in samples with GB *(A)* and radionecrotic *(B)* histologies.

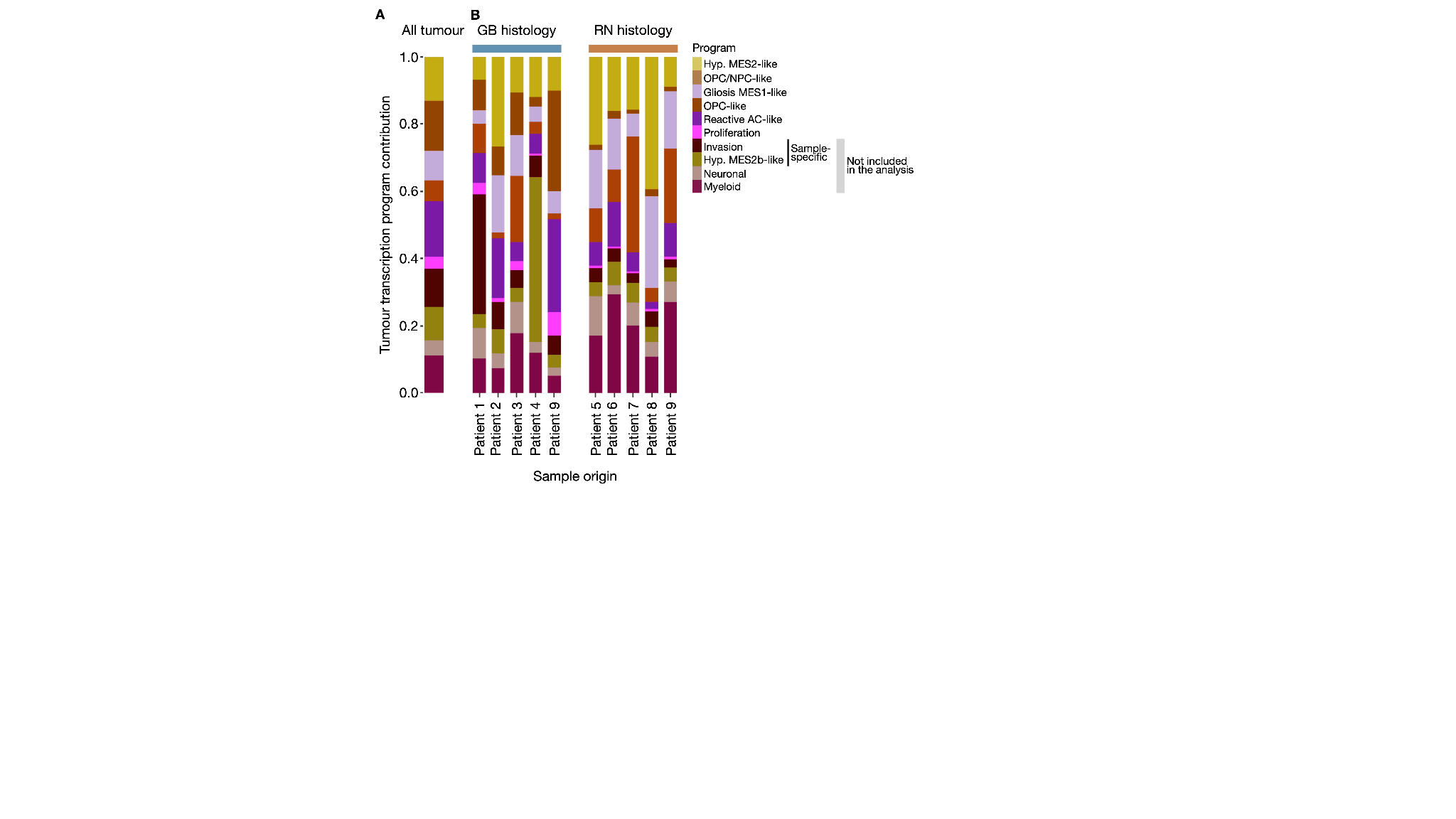

**Supplementary figure S5**. Signal contribution of tumour transcription programs for all tumour cells *(A)* and tumour cells within each sample *(B)*.

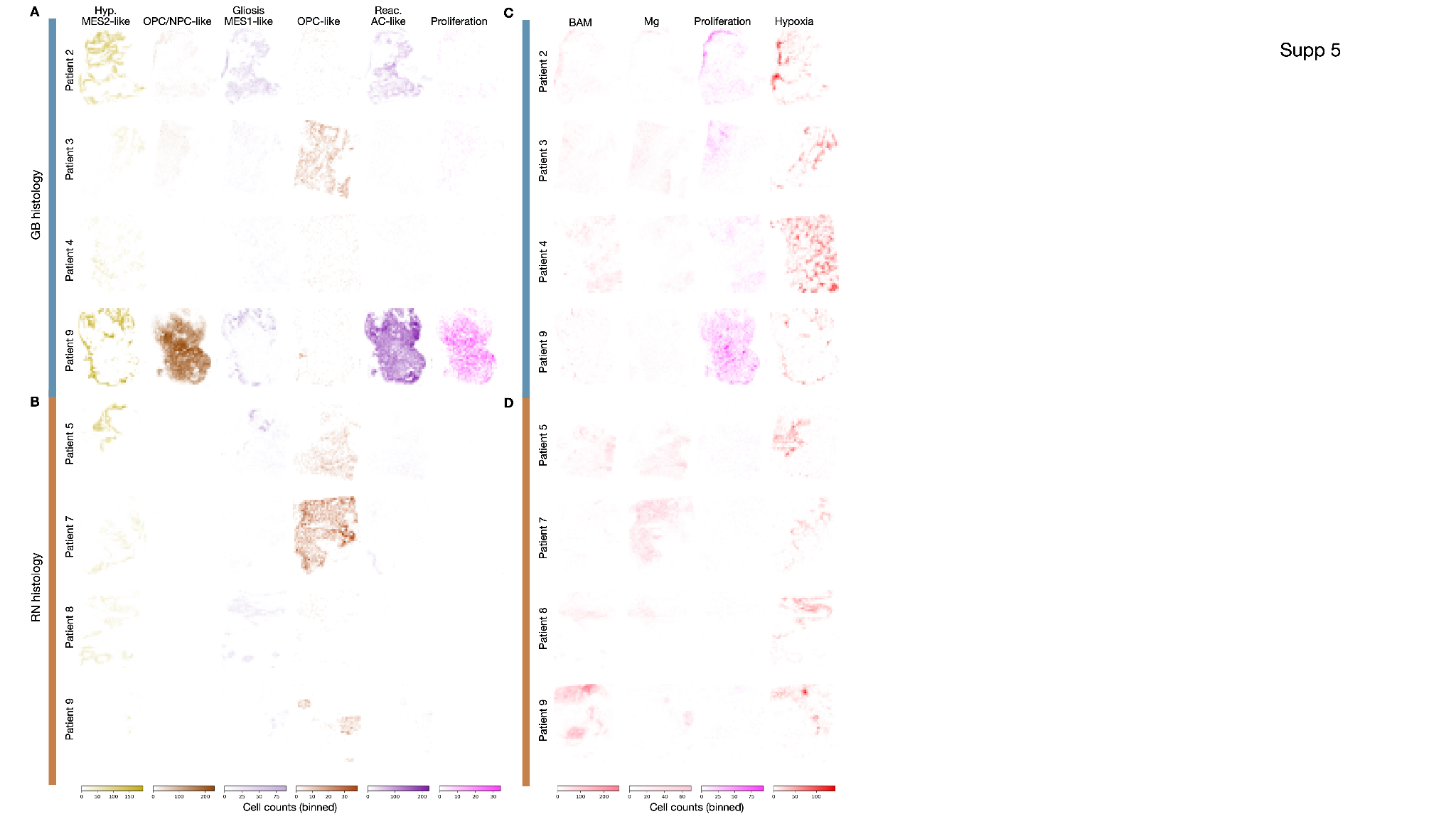

**Supplementary figure S6**. Spatial distribution of myeloid transcription programs in samples with GB *(A)* and radionecrotic *(B)* histology. Spatial distribution of myeloid transcription programs in samples with GB *(C)* and radionecrotic *(D)* histology.

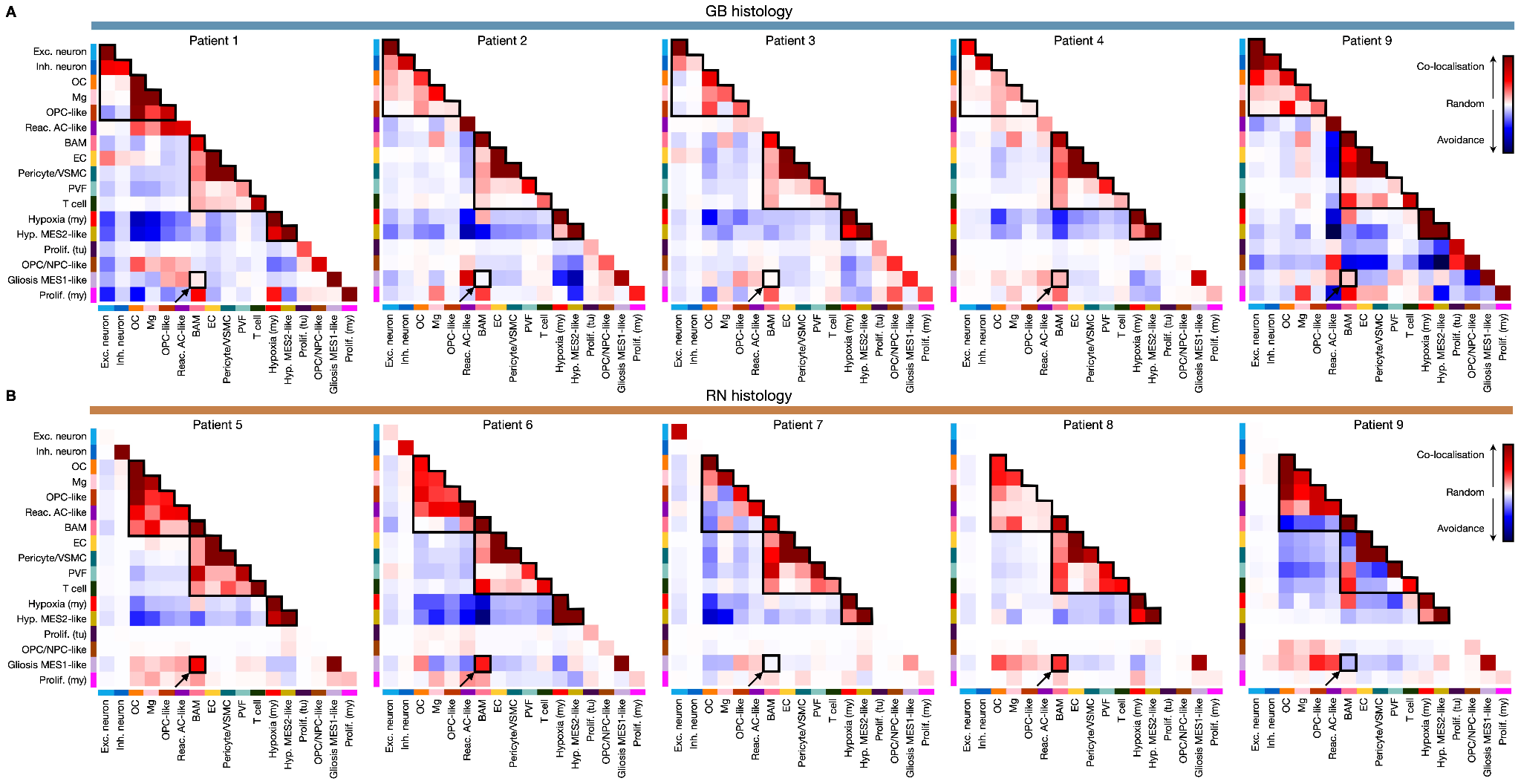

**Supplementary figure S7.** Cooccurring TME cell types and tumour programs form spatial communities in samples with GB *(A)* and radionecrotic *(B)* histology. My, myeloid transcription program; tu, tumour transcription program. Outlined spatial communities correspond to the three communities discussed in the main text. Arrows show BAMs colocalised with Gliosis MES1-like tumour cells.

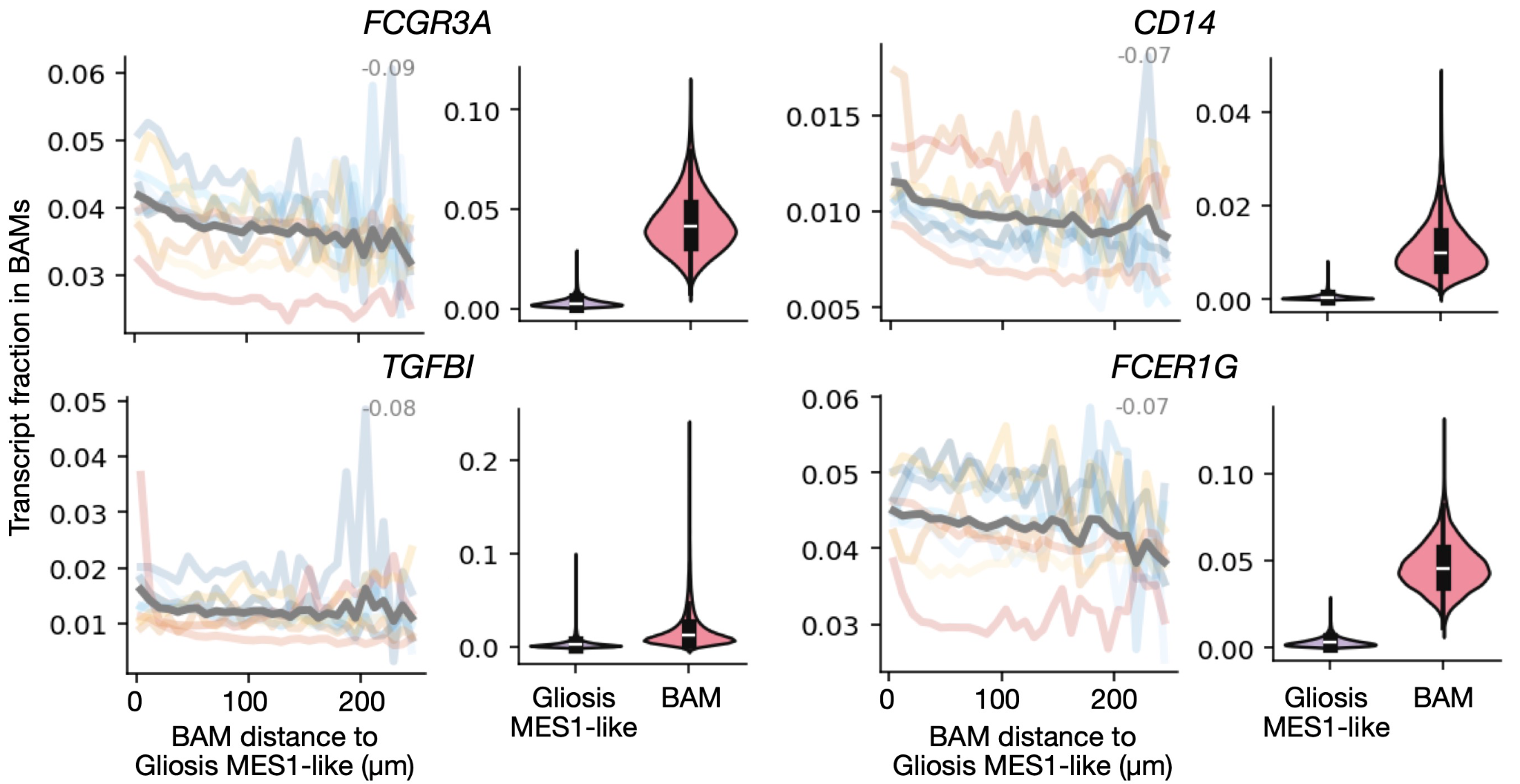

**Supplementary figure S8**. Selected genes with higher expression in BAMs close to Gliosis MES1-like tumour cells. Numbers correspond to Spearman correlation coefficient between BAM distance to the closest Gliosis MES1-like tumour cell and BAM gene expression.
